# Limited impact of lifting universal masks on SARS-COV-2 transmission in schools: The crucial role of outcome measurements

**DOI:** 10.1101/2023.08.26.23294658

**Authors:** Mingwei Li, Bingyi Yang, Benjamin Cowling

**Author notes:** **Corresponding author:** Bingyi Yang, School of Public Health, Li Ka Shing Faculty of Medicine, The University of Hong Kong, 7, Sassoon Road, Pokfulam, Hong Kong.

## Abstract

As the pandemic’s dynamics changed, many municipalities lifted face wearing requirement in school which was initially implemented to mitigate the transmission of COVID-19. This study examines the effects of lifting mask mandates on COVID-19 transmission within Massachusetts school districts. We first replicated previous research by Cowger et al. (2022) utilizing a Difference-in-Difference (DID) model. Then, we back project the case infection and calculate the Rt value to redo the DID analysis. However, when shifting the outcome measurement to the reproductive number (Rt), our findings suggest that lifting mask mandates can only significantly influence the Rt first two weeks post-intervention. This implies that while mask mandate plays a role in mitigation, its lifting does not drastically influence COVID-19 transmissibility in the long term.

## INTRODUCTION

Non-pharmaceutical interventions (NPIs) have played a critical role in reducing transmission during the COVID-19 pandemic and “flattening the curve”, spreading out infections over a longer period of time to reduce pressure on healthcare systems. One such NPI, recommending or even requiring people to wear face masks in certain settings or locations, was commonly used during the COVID-19 pandemic. Although challenges exist in examining the effectiveness of face masks in preventing disease transmissions that caused by the heterogeneity in mask materials, inconsistency in user compliance and the coincidental implementation of other NPIs, it was later suggested to be effective in mitigating COVID-19 transmissions and adopted by schools (Boutzoukas et al., 2022; Kim et al., 2020; Sharma et al., 2020). As the pandemic progresses, many local authorities relaxed mask mandates during periods when the pandemic appears to be receding, while the impact of lifting these mandates on transmission remains under-investigated. It’s critical to understand both the potential reduction in transmission when a mask mandate is introduced, as well as the possible increase in transmission when such interventions are relaxed or removed.

Cowger *et al*. found significantly lower incidence in schools which maintained universal masking compared to those without in the subsequent surge after mask mandates were lifted in Massachusetts school in February 2021 (Cowger et al., 2022), However, the outcome measurement (i.e., incidence) alone cannot fully reflect the transmission process since it does not account for the exponential changes in case numbers during the transmission. For instance, initial incidence discrepancies between two locations can magnify over time, even if the reproductive number remains constant. Similar oversights of transmission outcomes have been identified in studies about effectiveness of other COVID-19 control measures (Auger et al., 2020), which may have impacted the accuracy of the findings and their implications.

In this study, we used the same data source as Cowger *et al*. to examine the impact of outcome measure (i.e., transmission and/or incidence) on the estimated effectiveness of lifting masks mandates on COVID-19 transmission. After replicating the original analyses using a difference-in-difference model on incidence rate, we replaced the outcome measurement with the effective reproductive number (Rt). Rt is estimated as the average number of secondary infections resulting from one infected individual, measuring the transmissibility in a population (Nash et al., 2022). The Rt reductions are often used to evaluate the effectiveness of mitigation measures in limiting the virus transmission, with Rt below one indicating non-sustainable transmission under existing measures.

## RESULTS

We replicated Cowger et al. (2022) analysis using population-weighted COVID-19 incidence as the outcome measurement in a difference-in-difference model. We analysed data from 70 school districts in Massachusetts, with 46 lifting the mask mandates in the first reporting week (type 1), and 15 and 9 districts lifting in the second (type 2) and third (type 3) reporting week, respectively. Chelsea and Boston retained their mandates (type 4). We only replicated results for students, as data on school staff were not available.

Our replication results were consistent with those reported by Cowger et al.’s. We found that lifting the masking mandates in schools was associated with a notable increase in COVID-19 incidence across all district types (Figure 2A), with the lowest incidence in type 4 district that did not lifted the mask mandate. We observed an average treatment effect (ATT) of 39.1 (95% CI: 20.4-57.4) COVID-19 cases per 1,000 students associated with lifting masking mandates, compared to 39.9 (95% CI: 24.3-55.4) in the original analysis (Cowger et al., 2022). Analyses using incidence as outcomes suggested that lifting the masking mandates in schools was associated with 39.1 additional cases per 1,000 students within a 15-week period (Figure 2C).

**Figure 1.**
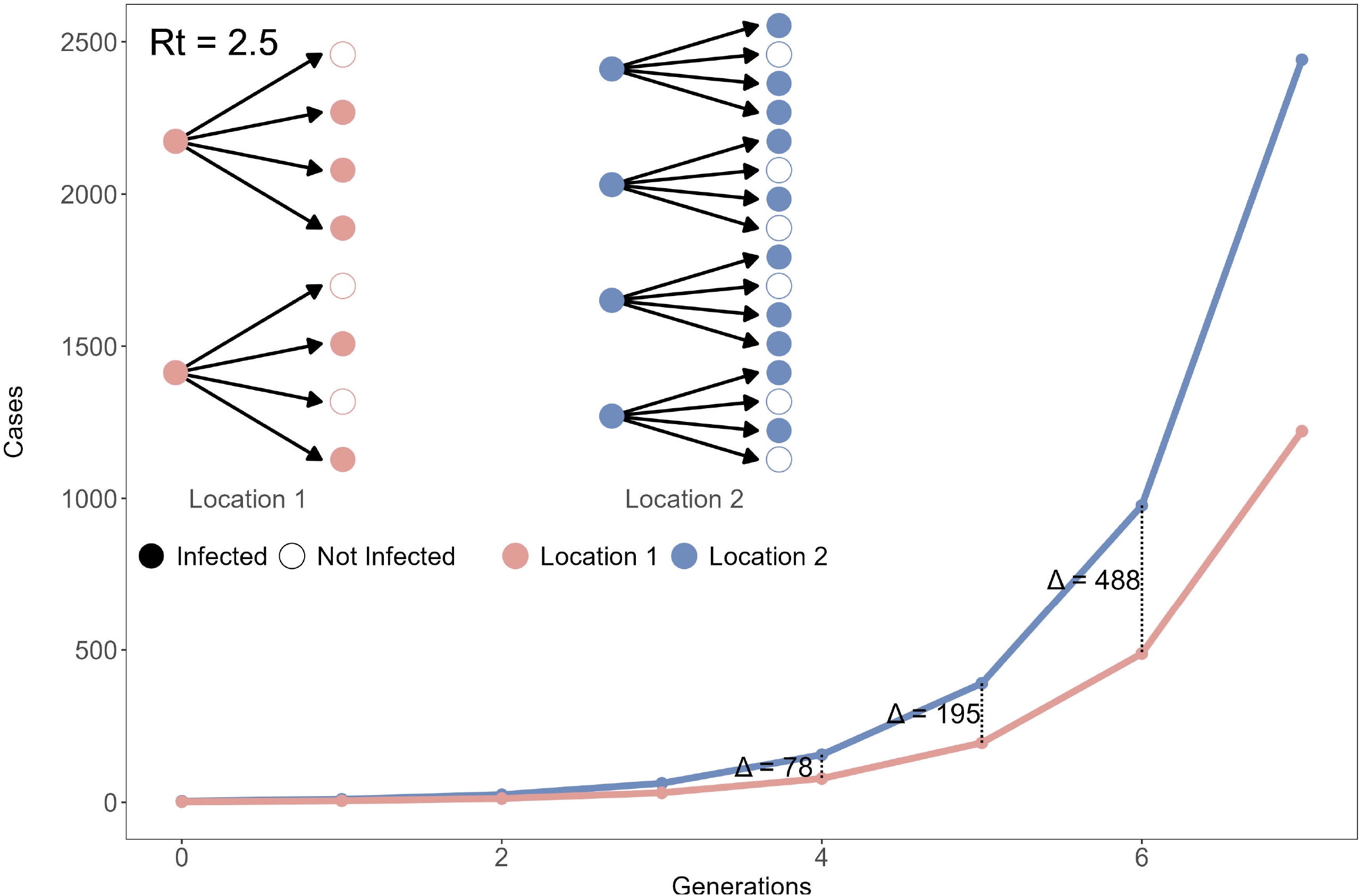
Hypothetical Demonstration of epidemic dynamic of 2 locations with identical Rt.

**Figure 2.**
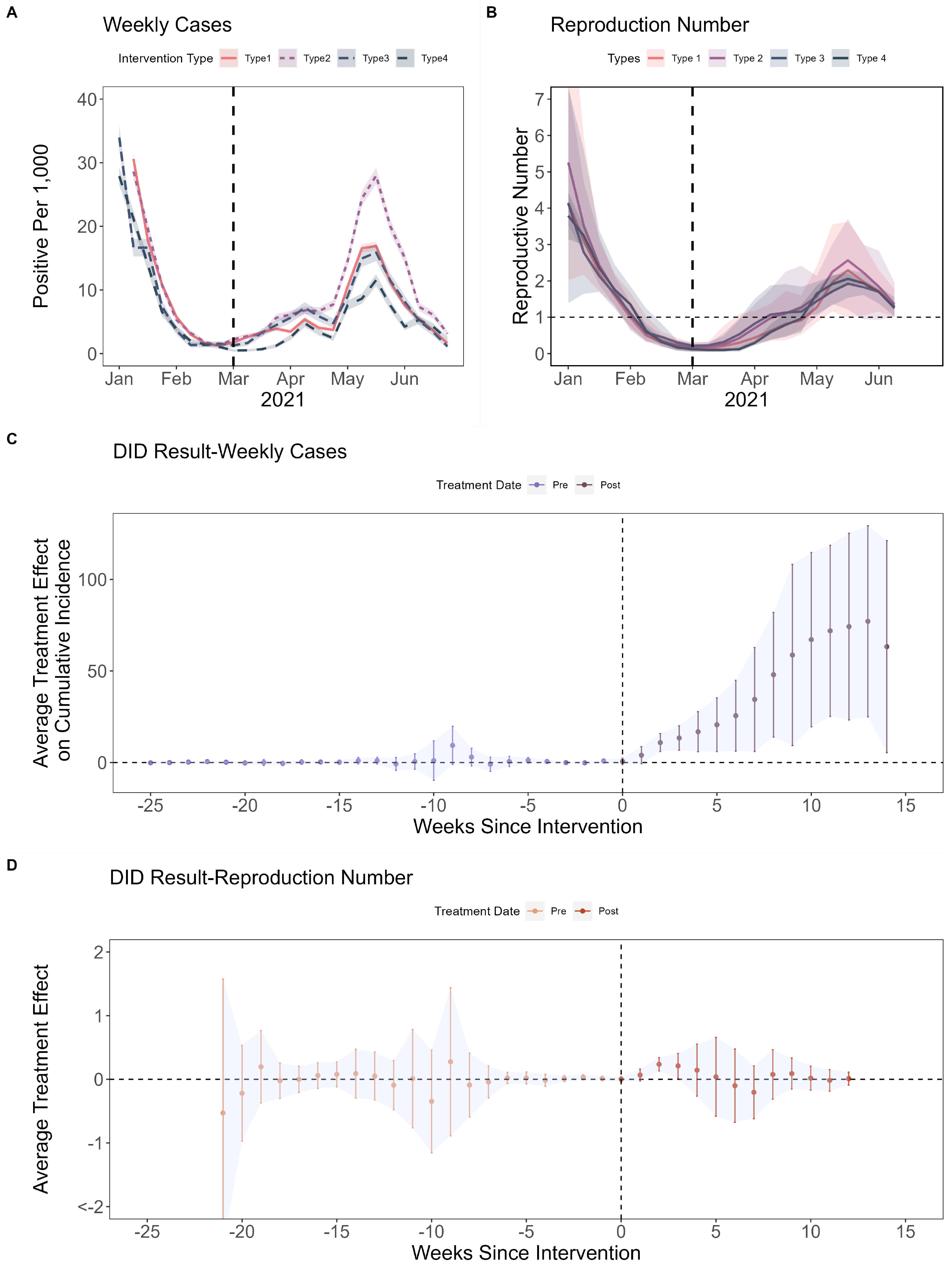
Difference-in-difference (DID) model analyses of COVID-19 incidence and effective reproductive number (Rt). **(A)** Weekly COVID-19 case trend by types of school district. **(B)** Daily COVID-19 Rt estimated from back projected incidence of students, stratified by types of school districts. **(C)** Average treatment effects (ATT) of the lifting masks mandates on cumulative incidence in students. **(D)** Average treatment effects (ATT) of the lifting mask mandates on Rt.

We further analysed the impact of lifting masks mandates on changes in Rt, which measures COVID-19 transmission intensity. Rt was estimated using method by Cori *et al*.’s method with a mean serial interval of 4.4 days (standard deviation 3.0 days) (Alene et al., 2021). Contrary to the original results using incidence rates, we found no significant difference in Rt across district types, except for type 4 district, which showed a slightly higher Rt in the first two weeks after lifting the mask mandates (Figure 2B). There was no association between lifting mask mandates and reductions in Rt (ATT 0.04, 95% CI, -0.11 to 0.20) throughout the entire post-lifting period (Figure 2D). Moreover, Rt remained below 1 from February to May, indicating a general decline in the epidemic during that time until a new wave of the pandemic in late April. (Figure 2B).

Sensitivity analysis yielded consistent results when using longer serial intervals (Bi et al., 2020; Xu et al., 2020) (Table S1) and weighting difference-in-difference model with populations (Brantly Callaway, 2022; Callaway & Sant’Anna, 2021) (Table S2). Our difference-in-difference analysis satisfied the parallel trend assumption and, according to Cowger *et al*. (2022), other time-varying variables including vaccination rate and community transmission remained stable during the study period.

## DISCUSSION

We found no evidence that lifting mask mandates in Massachusetts schools significantly affected COVID-19 transmission rates, which is contrary to the findings reported by Cowger et al. Our findings demonstrate that substantial changes in incidence or case numbers do not necessarily reflect substantial changes in underlying transmission. Additionally, we found non-sustainable transmission (i.e., Rt < 1) across all school districts before masks mandates were lifted, suggesting that factors other than lifting mask mandates impacted COVID-19 transmission in these schools, such as community transmission and the implementation of other measures (e.g., extensive testing) (Braga et al., 2022; Falk et al., 2021; Head et al., 2022; Juutinen et al., 2023).

Our findings highlight the importance of considering transmissibility outcomes when assessing the effectiveness of interventions against disease transmission(Cowger et al., 2022). While as counts-based outcomes (e.g., incidence) can serve as proxies for the difficult-to-measure transmission process, it is crucial to note that nonpharmaceutical interventions work by reducing person-to-person transmission, reducing subsequent incidence from what it would have been without the interventions. Due to exponential case growth and delays in disease development (e.g., incubation periods), changes in case counts may exaggerate and lag changes in transmission. Therefore, caution is necessary when interpreting the effects of interventions based on solely on analyses of incidence rates.

Our study has some limitations. Firstly, we were unable to account for unobserved heterogeneities, such as mask types and compliance with mask mandates. Therefore, our findings reflect the impact of the lifting of masks mandate policies rather than the direct effects of mask-wearing (Gettings et al., 2022; Tupper & Colijn, 2021). Secondly, we only estimated Rt using student cases, and we were unable to account for factors such as transmission between student and staff and external community due to data availability. Consequently, our results may partially capture the within-school transmission dynamics.

In summary, we found limited impact of lifting mask mandates on reducing COVID-19 transmission in Massachusetts schools in February 2021. Future assessments of the effects of interventions against transmission should consider including transmissibility outcomes.

## Data Availability

All data produced are available online at https://www.nejm.org/doi/10.1056/NEJMoa2211029.

## ACKNOWLEDGMENTS

This work was financially supported by a grant from the Research Grants Council of the Hong Kong Special Administrative Region, China (Project No. T11-705/21-N).

## AUTHOR CONTRIBUTIONS

All authors meet the ICMJE criteria for authorship. The study was conceived by ML, BY and BJC. Data analyses were done by ML. ML and BY wrote the first draft of the manuscript, and all authors provided critical review and revision of the text and approved the final version.

