## Supplementary Tables for "Limited impact of lifting universal masks on SARS-COV-2 transmission in schools: The crucial role of outcome measurements"

**Table S1. ATT using different sets of Serial Intervals**

| ***No Covariate*** |  |  |  |  |
| --- | --- | --- | --- | --- |
| ***SI (95% CI)*** | ***ATT*** | ***Std.Error*** | ***(Lower_95CI*** | ***Upper_95CI)*** |
| 4.4 (6.7 – 2.9) | 0.03 | 0.05 | (-0.07 | 0.14) |
| 5.3 (5.9 – 4.7) | -0.06 | 0.10 | (-0.14 | 0.26) |
| 6.3 (5.2 - 7.6) | 0.10 | 0.11 | (-0.11 | 0.32) |

**Table S2. ATT using different sets of Serial Intervals in a population weighted DID**

| ***Population Weighted*** | |  |  |  |
| --- | --- | --- | --- | --- |
| ***SI (95% CI)*** | ***ATT*** | ***Std.Error*** | ***(Lower_95CI*** | ***Upper_95CI)*** |
| 4.4 (6.7 – 2.9) | 0.00 | 0.03 | (-0.05 | 0.06) |
| 5.3 (5.9 – 4.7) | 0.01 | 0.04 | (0.06 | 0.08) |
| 6.3 (5.2 - 7.6) | 0.03 | 0.05 | (-0.08 | 0.13) |
